# Long Term Safety and Efficacy of Sub-Lingual Ketamine Troches / Lozenges in Chronic Non-Malignant Pain Management

**DOI:** 10.1101/2020.07.13.20153247

**Authors:** Ben Maudlin, Stephen B. Gibson, Arun Aggarwal

**Affiliations:** Pain Management Centre, Royal Prince Alfred Hospital, Missenden Road, Camperdown, NSW, Australia 2050

**Keywords:** Ketamine, lozenges, troches, chronic non-malignant pain

## Abstract

**Introduction:** Chronic non-malignant pain is a disabling condition that results in a reduction in function and quality of life when inadequately managed. Sub-lingual ketamine has been shown to be efficacious for use in chronic pain. Despite its use for decades in chronic non-malignant pain, there is no published long-term data on safety, side effects or adverse drug reactions. The aim of this case-series is to provide the initial evidence for safety and efficacy in this patient group.

**Methods:** We present a retrospective review of twenty-nine patients (n=29) from a metropolitan tertiary pain service who have been receiving sub-lingual ketamine troches / lozenges between the period of 2012-2019. Patients were identified from the outpatient pain clinic, who had been admitted for inpatient subcutaneous ketamine infusions as part of opiate detoxification or management of central sensitisation due to a chronic neuropathic pain syndrome. An initial review was performed to check the patient started taking the ketamine troches. Each of these medical records was reviewed manually to extract information to a datasheet.

**Results:** There was a wide range of dosages used from 25-600mg in divided doses. The duration of treatment ranged from 2-89 months. There was no association with either the dosage or duration of treatment and frequency of side effects. There was an overall reduction in the use of opioids, gabapentinoids or benzodiazepines in 59% of patients with 39% having a complete cessation of an analgesic agent. Side effects were reported in 24%, but only 7% discontinued the treatment due to the side effect (drowsiness). There were no reports of renal impairment, cystitis, or hepatotoxicity.

**Discussion:** This retrospective case-series has demonstrated that sub-lingual ketamine is a safe and effective analgesic agent to use in chronic non-malignant pain management. It is indicated in a variety of chronic pain conditions and has an excellent safety profile, with no association between the frequency in side effects and duration of therapy or total daily dosages. The study has also shown that the “safe” dose may be higher than the previous consensus.

**Contribution Statement:** A.A and S.G. recruited the patients. A.A. & B.M. created the parameters for the data collection sheet. B.M Collected most of the data from the medical records, entered it into a datasheet, wrote the manuscript, ran the statistics, performed the data analysis, and generated the figures and tables. A.A. Edited the manuscript. S.G. and A.A were the research supervisors.

## INTRODUCTION

Chronic pain is a complex and challenging illness with significant impacts on both patients and their families and has a significant social and economic burden. It has been demonstrated that sub-lingual ketamine is efficacious for use in chronic pain, particularly for opioid-induced hyperalgesia (OIH), chronic neuropathic pain and complex regional pain syndrome (CRPS) [13; 20; 29].

Ketamine acts on both N-methyl-D-aspartate receptor (NMDA) and non-NMDA receptors. It reduces the sensitisation of the central nervous system (CNS) to painful stimuli. It has the benefits of reducing the numerical pain rating scale (NPRS) and, in select cases, an opioid-sparing effect [18; 27]. Sublingual administration has a bioavailability advantage over oral administration.

Reported adverse effects in the literature include psychotomimetic effects (hallucinations, memory deficits, dysphoria), nausea and vomiting, cardiovascular stimulation [23]. When abused recreationally (at significantly higher doses than used in chronic pain), there is an increased frequency of cystitis, urinary frequency, cognitive disturbances, and theoretical neurotoxicity (in experimental models only) [12; 17].

However, there currently is no published long term safety data on ketamine [3; 4]. This case series aims to provide the initial evidence for the safety and efficacy of oral ketamine in this patient population with a secondary aim in examining the reduction in other analgesics due to the introduction of sub-lingual ketamine into the regimen. We hope this will provide the needed safety data for further studies that will provide the robust data needed to received PBS approval for this indication.

### Pharmacology

Ketamine is a non-competitive N-methyl-D-aspartate receptor antagonist (NMDA); it also exhibits effects on non-NMDA receptors (opioid, monoaminergic, cholinergic, nicotinic, and muscarinic receptors). It has three main effects, analgesic, dissociative/hypnotic and sympathomimetic. Its analgesic effects come primarily from the inhibition of afferent signals from painful stimuli via the spino-reticular pathway. It achieves this primarily by binding to the NMDA receptor (specifically NR2D) intraductal PCP site, decreasing the channel opening time [21; 23]. Ketamine decreases the response to a repeated stimulation or “wind up” process of sensitising the CNS to pain [2; 14]. These NRD2 receptors are primarily responsible for nociception and almost exclusively located in the spinal cord. This action is also why there is benefit in OIH, where glutamate fixation and activation of NMDA receptors leads to crosstalk of neural mechanisms of pain and tolerance [2; 11; 14].

Ketamine undergoes extensive metabolism, initially to norketamine, primarily via Cytochrome P450, norketamine is further metabolised to the hydroxynorketamine both these metabolites are both active and antagonise the NMDA receptor.

It undergoes extensive first-pass metabolism when administered orally. The bioavailability of ketamine is dependent on the route in which it is administered, with a bioavailability of between 7-9% orally (when swallowed) and 17-29% sublingually with a terminal half-life of 183 minutes; this is sufficiently high and reproducible to support its use in pain management [6–8; 26; 33]. The sublingual route is advantageous as it has reduced first-pass metabolism; however, the active metabolites’ plasma concentrations are similar in both sublingual and oral routes [26; 33].

High plasma concentrations are required to achieve dissociative (50-100ug/L) and hypnotic effects (>2000ug/L). Ketamine preferentially inhibits excitatory-to-excitatory coupling above inhibitory-to-excitatory, as is the case with most anaesthetic agents that potentiate or are GABA agonists [30]. The sympathomimetic properties are mediated by enhanced central and peripheral monoaminergic transmission (dopamine and noradrenaline) [1]. Ketamine has the additional benefit of enhancing the descending inhibiting serotoninergic pathway and inhibiting serotonin uptake and thereby exerts anti-depressive effects, unlike any other anaesthetic agents, this may play a vital role in the emotional response to pain perception [19; 22]. However, it is difficult to separate the confounding effects of a reduction in pain on mood from antidepressant activity or attenuation.

## METHODS

This retrospective case series identified twenty-nine patients (n=29) from a metropolitan tertiary pain service who received sub-lingual ketamine troches/lozenges between 2012-2019. Patients were identified from a tertiary multi-disciplinary outpatient pain clinic who had already undergone a comprehensive multi-disciplinary pain management assessment. They were admitted for inpatient subcutaneous ketamine infusions as part of opioid detoxification or central sensitisation management due to chronic non-malignant pain. The doses received varied from patient to patient. They all received a continuous subcutaneous infusion at 0.1 to 1.2 mg/kg/hour (maximum daily dose: 500 mg) titrated to pain and tolerability [10]. Only patients who responded to ketamine infusions went on to receive ketamine troches. The response was defined as a >20-point reduction on a 0 to 100 mm visual analogue scale (VAS). After the initial infusion, there were no further subcutaneous infusions given.

The initial dose was commenced at a low dose of 25mg three times daily and titrated to tolerability and effect. Patients were seen in the pain clinic on a three-monthly basis to review and provide schedule 8 Special Access scheme (S8 SAS) scripts. Community compounding chemists compounded the troches/lozenges via the SAS scheme as it is off-label drug use. Troches were dispensed monthly as most S8 medications. An initial review was performed to check the patient started taking the ketamine troches. Each of these medical records was reviewed manually for the onset of symptoms; chronic pain diagnosis; commencement of sub-lingual ketamine; duration of therapy; cessation reason; adverse reactions or adverse effects; any opioid or other analgesic reduction; and numerical pain rating scale pre and post-commencement of sub-lingual ketamine. Sublingual ketamine was used for its advantageous bioavailability.

Underlying diagnoses were made or confirmed by Consultant Pain Specialists using accepted or suggested diagnostic criteria/classifications, including those developed by the American Pain Society (APS) and American College of Rheumatology (ACR) and notably the International Association of the Study of Pain (IASP) classification of chronic pain for ICD-11: chronic primary pain [25]

### Data Analysis

The Sydney Local Health District Human Ethics Committee approved the study. The data points that were examined and compared were gender, age, pain scores, total daily dose, duration of pain before initiation of oral therapy and duration of oral ketamine therapy. We calculated descriptive statistics and 95% confidence intervals (95% CIs) using statistical software (The jamovi project (2020). jamovi (Version 1.2) [Computer Software]. Retrieved from https://www.jamovi.org). When numerators were zero, we calculated 95% CIs using the methodology of Hanley and Lippman-Hand [15]. Means and standard deviations are reported, normal distribution of data allowed parametric analysis to be performed. A posthoc analysis was performed using a Spearman Rank Correlation (*r*_*xy*_) and determined in the statistical software using a correlation matrix based on R Core Team (2018). R: A Language and environment for statistical computing. [Computer software]. Retrieved from https://cran.r-project.org/. This was chosen as it evaluates the monotonic relationship between the ranked values. In a monotonic relationship, the variables tend to change together, but not necessarily at a constant rate. As per statistical protocol, a p-value <0.05 was considered statistically significant.

### Demographics

Background data is presented descriptively with means, standard deviations and comparisons between the sexes (Table 1). Descriptive statistics (means and SDs) of the outcome variables were computed for the sample and were grouped according to sex. Each was first reported using descriptive statistics, and then comparisons were carried out between the sexes for each instrument using Chi-Squared and t-tests (two-tailed). Our analysis identified that gender does not impact the safety and tolerability of ketamine.

**Table 1:** Differences between men and women regarding their age, duration of pain, the total daily dose of ketamine, duration of treatment, and pain score reduction (NPRS).

|  | *All*<br>(*n*=29) |  | *Women*<br>(*n*=19) |  | *Men*<br>(*n*=10) |  | *Sig* |
| --- | --- | --- | --- | --- | --- | --- | --- |
|  | Mean | SD | Mean | SD | Mean | SD | *p* |
| *Age* | 48.0 | 13.9 | 51.1 | 15.5 | 42.3 | 7.9 | 0.055 |
| *Duration of Pain (months)* | 177.1 | 147.38 | 205.6 | 174.38 | 126 | 56.64 | 0.089 |
| *Total Daily Dose (mg)* | 216.4 | 136.69 | 231.4 | 159.0 | 189.5 | 83.9 | 0.37 |
| *Duration of Treatment (months)* | 31.3 | 147.38 | 27.4 | 23.8 | 38.4 | 33.6 | 0.38 |
| *Pain Score Reduction (NPRS)* | 5.8 | 2.09 | 6.1 | 2.1 | 5.2 | 2.0 | 0.31 |

## RESULTS

The patients were a heterogeneous group. There were more women and men (19 vs 10), age groups had a wide variation of 26-67 years. There was no statistically significant impact of gender on measured outcomes. The diagnoses included peripheral neuropathy, complex regional pain syndrome, chronic neuropathic pain, trigeminal neuralgia, fibromyalgia, spinal syrinx, spina bifida and cauda equina (Table 2).

**Table 2:**
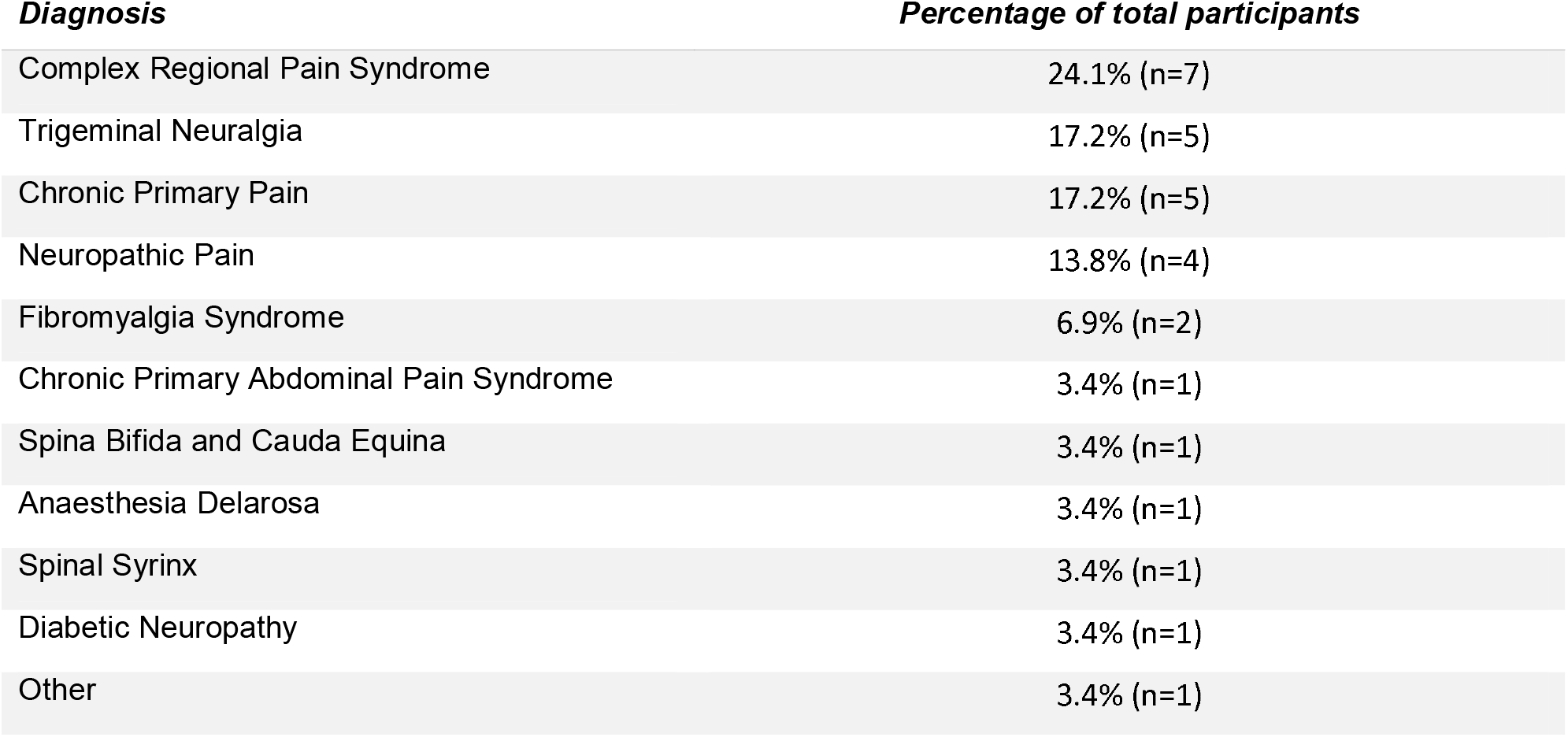
Patient Diagnosis as a percentage of total participants. Indications: Chronic non-malignant pain conditions that have responded poorly to conventional analgesics, opioids and anti-neuropathic medications, opioid tolerance, or opioid-induced hyperalgesia.

Before initiation of oral therapy, the duration of pain ranged from 24-600 months, with a mean of 177 months and a median of 126 months. There was no association between duration of pain and reduction in NPRS. The total daily ketamine dosage also varied from 25-600mg in three divided doses, with a mean dose of 216mg per day and a median of 200mg per day.

The reduction in NPRS ranged from 2-10 out of 10. The mean pain score before commencing any ketamine therapy was 8.71, and after ketamine was 2.89. (p<0.005), with a mean pain reduction of 5.82. There was a positive correlation (*r*_*xy*_ = 0.4305) between the total daily dose and the NPRS reduction from baseline to time of data collection, which was statistically significant (p=0.021). There was no correlation between total daily dose and frequency of adverse effects reported (*r*_*xy*_=0.3610, p=0.973).

The duration of treatment ranged from 2-89 months, with 59% (n=17) of patients having ongoing use. There was no association between the duration of use and the frequency of adverse effects, indicating that remaining on sub-lingual ketamine does not increase adverse effects. Also, there was no association between the duration of use and the change in NPRS, indicating a sustained response to sub-lingual ketamine.

The principal reason for discontinuation was the inability to obtain ongoing Department of Health authority (n=4) to approve sub-lingual ketamine in 2019 when the approving body was concerned about the lack of evidence for the long-term use of ketamine. The other reasons for discontinuation were the inability to afford the therapy (n=1); currently, in Australia, ketamine troches are not available on pharmaceutical benefits scheme at an approximate cost of $ 200/month, drowsiness (n=2), loss of efficacy (n=2), wanting to conceive a child (n=1) and insertion of a spinal cord stimulator (n=1) and not specified (n=3).

There were reductions in the use of opioids, gabapentinoids (GBN) or benzodiazepines (BZD) in 59% (n=17) of patients, with 39% (n=9) having a complete cessation of an analgesic agent (opioids, GBN or BZD).

### Adverse Effects

There were no serious adverse events (SAE). SAE only covers severe and life-threatening adverse effects such as death, danger to life, hospital admission, prolonged hospitalisation, absence from productive activity, increased investigational or treatment costs, and congenital disabilities. Adverse effects were self-reported; any reported adverse effects that occurred throughout the duration of therapy were documented. Of the patient group 24% (n=7) reported any adverse effects, 15% (n=5) reported excessive daytime drowsiness at doses sufficient to gain an analgesic effect, 9% (n=3) reported light-headedness/ dizziness, 3% (n=1) reported nightmares, and 3% (n=1) reported dysphoria, which resolved with a dose reduction.

Of these seven patients with adverse effects, only two discontinued the treatment due to the adverse effect (drowsiness). There were no reports of renal impairment, cystitis, or hepatotoxicity. This is of particular note as previous studies have indicated these adverse effects in their patient groups [3]. All patients were followed up for at least 31 months after the initiation of therapy. Previous studies indicated that renal impairment and ulcerative cystitis this would occur within the first 24 months and were mainly associated with recreational ketamine doses of more than 1000mg daily [24; 31; 32].

One patient attempted suicide and used their troches as an analgesic agent to assist their physical suicide attempt; the therapy was subsequently discontinued. There was no correlation between the total daily dose (r=0.369, p=0.973), duration of therapy (r=0.051, p=0.603) and adverse effects, however it was not adequately powered due to the small sample size and therefore there is a risk of a type II error.

## DISCUSSION

Ketamine has been shown to reduce excessively painful responses by antagonising NMDA receptors, improving opioid receptor sensitivity, reducing opioid tolerance, and suppressing opioid-induced hyperalgesia, resulting in a reduced central sensitisation component of pain [16]. Several studies that reported successful short-term treatment of chronic non-malignant pain with a ketamine infusion [28] Long-term efficacy of sub-anaesthetic, sub-cutaneous ketamine in the setting of chronic non-malignant pain has also been demonstrated [34]. This retrospective study analysed whether subsequent treatment with sub-lingual ketamine troches/lozenges resulted in longer-term efficacy of the beneficial effects of the initial ketamine infusion and assessed the long-term safety.

Oral or sub-lingual ketamine formulations are not currently commercially available but can be formulated by compounding pharmacists. Studies have shown that the sublingual formulation’s bioavailability is superior to an oral formulation, 25% compared to 10% [6–8]. There is substantial metabolism to nor-ketamine, which possesses analgesic activity, from both routes. The mean nor-ketamine / ketamine area under the plasma concentration-time curve from baseline to 8 hours ratios were 5 and 2.1 after sublingual and oral administration, respectively [23].

The patients from our practice typically offered an inpatient sub-anaesthetic, sub-cutaneous ketamine infusion are those who have failed a wide range of pharmacological and cognitive behavioural therapy options, noting that all are co-managed within our service by both a pain physician and a mental health practitioner, either a psychiatrist or clinical psychologist.

Therefore, the overall reduction in the use of opioids, gabapentinoids or benzodiazepines in 59% of patients after the inpatient subcutaneous ketamine infusion with 39% having a complete cessation of an analgesic agent as a result of outpatient therapy with ketamine troches, which is relatively impressive given the difficulties faced in treating this subset of patients. The relative dose of the ketamine troches is small compared to the amount needed for recreational use/abuse. However, the risk of misuse is still possible but given the potential benefit of this therapy, clinician vigilance is recommended, as with all drugs of potential abuse.

Of the patient group, 24% reported any adverse effects, but only 7% discontinued the treatment due to the adverse effect (drowsiness). There were no reports of renal impairment, cystitis, or hepatotoxicity.

The total daily dosage also varied from 25-600mg in divided doses, with a mean dose of 216mg per day and a median of 200mg per day. This dose is higher than the previously considered “safe” daily dose of 150mg per day [9].

The treatment duration ranged from 2-89 months, with 59% (n=17) of patients having ongoing use of ketamine troches at the time of study completion. The mean duration of treatment was 31.3 months. There was no association between the duration of use and the frequency of adverse effects indicating that remaining on sub-lingual ketamine long-term does not increase adverse effects.

## LIMITATIONS

We accept that our study has limitations, as it is a retrospective, non-randomised, non-blinded study. Adverse events were self-reported. This would ideally be collected in a questionnaire at each visit. This has subsequently been introduced at our centre. The NPRS is subjective, influenced by many factors that cannot be measured precisely, like mood, cultural factors and education. The recording of other measures, such as quality of life scores and depression scores, would have been helpful to interpret changes in NPRS scores in individuals further, but this was beyond the scope of this observational study. The data was taken from medical records that rely on the clinician to document accurately, and there is a risk of omission and transcription error.

In a prospective study, the troches would be compounded centrally, as this was a retrospective series and had not occurred before data collection. Given that ketamine is an accountable and controlled drug, two pharmacists would have verified the amount of ketamine during the compounding process. Ideally, plasma levels would have been collected to ensure the troches’ absorption and regular monitoring of LFTs and urine samples; this had not occurred for this data set.

We acknowledge that this is a relatively small study. However, given that there is a paucity of information regarding the long-term effectiveness of sub-lingual ketamine currently in the literature, even this small study provides valuable information regarding this under-utilised treatment to manage chronic non-malignant pain. Given the positive results of this study, we are currently undertaking a more extensive study to provide additional information on the safety of sub-lingual ketamine to treat chronic non-malignant pain.

## CONCLUSION

This retrospective case series has demonstrated that sub-lingual ketamine is safe and provides a substantial reduction in pain scores and other analgesic agents to use in chronic non-malignant pain management. It is indicated in various chronic pain conditions and has an excellent safety profile, with no association between the frequency of adverse effects and duration of therapy or total daily dosages. The study has also shown that the “safe” dose may be higher than the previously suggested maximum of 1.5mg/kg per dose or 5mg/kg/day [3; 5].

## Data Availability

Data was collected from Pain Clinic Letters and Electronic Medical Records

## Device status

None declared

## Ethics

The Sydney Local Health District Human Ethics Committee approved the study. It was deemed a low-negligible risk study.

